# Dietary Supplement Promotion by Social Media Influencers: Risk Omission and Limited Disclosure

**DOI:** 10.64898/2026.10.27.26366588

**Authors:** Elena Engel, Kathrin Karsay, Michelle Möri, Isa Vos, Thomas Friemel, Brooke Nickel, Raffael Heiss

**Affiliations:** University of Vienna, Vienna, Austria; MCI Management Center Innsbruck, Innsbruck, Austria; University of Zurich, Zurich, Switzerland; The University of Sydney, Sydney, Australia

## Abstract

**Importance:** Dietary supplements are commonly used by adolescents and can carry risks including adverse effects, drug interactions, and contamination-related harms. Social media influencers are prominent sources of health, fitness, and nutrition information for adolescents, yet little is known about how supplements are promoted in influencer environments relevant to young people or whether potential risks are communicated alongside claimed benefits.

**Objective:** To characterize dietary supplement promotion among Instagram influencers identified through audience-centered sampling in studies of adolescents, focusing on supplement types, advertising disclosure, persuasive strategies, and communication of benefits and harms.

**Design, Setting, and Participants:** Cross-sectional content analysis of 1120 dietary supplement presentations in 727 Instagram posts by 84 influencer accounts identified through 2 independent youth-focused studies in Switzerland and Austria.

**Main Outcomes and Measures:** Main outcomes were supplement types, overall tone, advertising disclosure, persuasive strategies, and communication of benefits and harms. Mixed-effects logistic regression models with random intercepts for influencer accounts examined associations of influencer characteristics and supplement categories with personal experience narratives and benefit claims. Outcomes with sparse distributions were summarized descriptively.

**Results:** Of 1120 supplement presentations, 1092 (97.5%) were promotional in tone and 1088 (97.1%) involved advertising, of which 802 (73.7%) lacked disclosure. Sports nutrition supplements accounted for 566 presentations (50.5%). Personal experience narratives were present in 458 presentations (40.9%), whereas evidence cues were present in only 42 (3.8%). Benefits were mentioned in 269 presentations (24.0%), approximately 10 times as often as harms (26 [2.3%]), and no presentation mentioned drug interactions or contamination risks.

**Conclusions and Relevance:** Among influencers identified through audience-centered sampling in studies of adolescents, dietary supplement promotion was highly commercialized, advertising was frequently undisclosed, and benefit communication substantially outweighed risk communication. The scarcity of harm communication and omission of clinically relevant risk information suggest that influencer environments relevant to adolescents may provide limited information for evaluating the potential risks of dietary supplements.

**Key Points:** *Question:* How are dietary supplements promoted by social media influencers identified through youth-focused sampling, and to what extent do they communicate risks alongside claimed benefits?

*Findings:* In this cross-sectional content analysis of 1120 dietary supplement presentations across 727 Instagram posts by 84 influencers identified through 2 youth-focused studies, 97.1% involved advertising, of which 73.7% lacked disclosure. Benefits were mentioned approximately 10 times as often as harms, and no presentation mentioned drug interactions or contamination risks.

*Meaning:* Dietary supplement promotion in influencer environments relevant to adolescents is highly commercialized, frequently undisclosed, and provides limited clinically relevant safety information.

## Introduction

Dietary supplements are widely consumed across age groups and are often perceived as safe and health-promoting.^1,2^ However, many products carry well-documented risks, including adverse effects, contamination, and interactions with prescription medications.^2–5^ Unlike pharmaceuticals, most supplements require no premarket demonstration of efficacy or safety, and regulatory oversight of manufacturing quality remains limited in most jurisdictions.^6^ These concerns are particularly relevant for adolescents and young adults, among whom supplement use is common and may be shaped by fitness, appearance, and performance goals.^7^

Increasingly, dietary supplements are presented through social media influencers who embed product endorsements within lifestyle, fitness, health, and wellness content.^8–11^ For adolescents, these influencers are prominent and relatable sources of health information.^7,8^ This raises concerns because many influencers lack formal health-related education and present supplements within commercial partnerships or affiliate marketing arrangements.^12,13^ Because promotional messages are often embedded in personal narratives and everyday lifestyle content, the distinction between advertising, recommendation, and personal experience may be difficult to recognize.^13,14^

Unlike health professionals, influencers are not bound by professional standards for balanced risk communication and clinical accuracy, while their endorsements may derive credibility from perceived authenticity and relatability.^12^ Prior research on influencer promotion of prescription drugs^13,14^ and medical tests^15,16^ has documented substantial imbalances in benefit and risk communication, with risk information frequently minimized or omitted. However, systematic evidence on dietary supplement promotion remains limited. Little is known about which ingredients are most frequently promoted, which persuasive strategies are used, the extent to which commercial relationships are disclosed, and whether potential harms are communicated alongside claimed benefits. This gap may be particularly consequential in influencer environments relevant to adolescents.

To address these gaps, we conducted a quantitative content analysis of dietary supplement presentations on Instagram using an audience-centered sampling approach that identified influencers through 2 independent studies of adolescents. By drawing on both adolescents’ reported encounters with social media accounts and participatory sampling by adolescents themselves, this approach was designed to capture influencers relevant to young people’s everyday social media environments. We examined supplement types, persuasive strategies, disclosure practices, and the communication of benefits and harms.

## Methods

### Study Design

We conducted a quantitative content analysis of publicly available Instagram posts presenting dietary supplements. The study protocol was preregistered on the Open Science Framework (https://osf.io/rw9pz). Ethics approval was obtained from the Ethics Board of the MCI – Management Center Innsbruck (No. 20250310). The coding scheme was developed using a combined deductive-inductive approach informed by prior literature and refined during pretesting (full codebook with prior literature in **eMethods 1**).

### Data Collection

Sampling proceeded in three stages. First, influencer accounts were identified using an audience-centered sampling strategy drawing on 2 independent youth-focused studies in Switzerland and Austria that used complementary approaches to identify accounts relevant to adolescents (**eMethods 2**). In the Swiss study, 1392 adolescents aged 14 to 18 years from 12 secondary and vocational schools were surveyed about social media content promoting healthier lifestyles. Adolescents who reported seeing such content were asked to nominate specific accounts from which they frequently encountered these messages. In the Austrian participatory study, 336 adolescents aged 12 to 19 years from 16 schools actively identified influencers they considered relevant within their social media environments and sampled health-related content from these accounts.^17,18^

This approach identified influencers based on their relevance to adolescent audiences rather than researcher-defined lists of health or supplement influencers. Between August and September 2024, the 2 datasets were merged, duplicates were removed, non-Instagram accounts were excluded, and accounts were screened for health-related content, including dietary supplements.

Second, between April and September 2025, eligible accounts were required to have at least 5000 followers and at least 5 supplement-related posts among their 100 most recent posts. This yielded 107 influencer accounts comprising 1572 posts referencing dietary supplements. Third, because the present analysis focused on specific products, only posts containing at least 1 identifiable product were retained, yielding 734 posts and 1134 supplement presentations from 86 influencers. During full content analysis, 14 supplement presentations spanning 7 posts across 3 influencer accounts were subsequently excluded because the underlying posts were no longer accessible online. Two of these accounts had no remaining eligible posts and were fully excluded, resulting in a final analytic sample of 1120 supplement presentations across 727 posts from 84 influencers.

### Coder Training and Reliability

The first author (EE) and two trained student coders (JS, KF) pretested the codebook on a subsample of the dataset (*n* = 42 cases), meeting after each round to review disagreements, clarify coding rules, and resolve ambiguities; reliability was assessed using Krippendorff α, with percentage agreement reported for variables where low prevalence precluded meaningful α estimation (**eMethods 3a**). The student coders then completed coding of the full dataset. For variables that did not reach adequate reliability (α < 0.80) during pretesting, two coders (EE, KF) recoded a subsample of the final dataset (n = 113 cases) using Cohen κ (**eMethods 3b**). All variables retained for the primary analyses demonstrated α or κ values of 0.80 or greater; for rare categories for which chance-corrected reliability could not be estimated, percentage agreement was reported.

### Measures

Influencer characteristics included gender, follower count, age, verification status, country of origin, and professional background (full measures in **eMethods 1)**. Influencers were classified as health professionals if they explicitly indicated a recognized health-related credential in their profile (eg, physician, registered dietitian, pharmacist, physiotherapist, or nurse); all others were classified as nonprofessional.

Identified supplement products were coded according to ingredients (eg, caffeine, protein, amino acids) and product form (eg, liquid, pills, gummies). Ingredients were further grouped into five supplement categories: (1) sports nutrition (protein, amino acids, creatine), (2) stimulants (caffeine), (3) micronutrients (multivitamins, multiminerals), (4) miscellaneous (herbal substances, collagen, omega-3/6 fatty acids, pre- and probiotics, antioxidants, melatonin, testosterone boosters, and other ingredients not otherwise classified), and (5) no specific ingredient identified. Because products could contain multiple ingredient types, supplement categories were not mutually exclusive.^19,20^

Persuasive strategies were coded using a predefined scheme capturing personal experience narratives, evidence cues (eg, references to studies or scientific language), and behavioral modeling, defined as the influencer’s visual interaction with the product (holding or presenting without consuming, using the product in a recipe without consuming the end product, or consuming the product or recipe containing it).^12,16,21^

Advertising was coded as present when a financial interest underlay the product mention. Presentations without financial interest, such as negative discussion in a reaction video, were not classified as advertising. For presentations classified as advertising, disclosure was subsequently assessed and coded as present if any verbal, visual, or textual indicator of a commercial relationship was included (eg, “ad,” “paid partnership,” or platform-native disclosure tools); tagging the brand or product alone was not considered sufficient disclosure.

Overall tone was coded as promotional/positive, negative/warning, or neutral/balanced; presentations identified as advertisements were coded as promotional unless framing explicitly emphasized potential harms over benefits.

Benefit framing was coded across multiple domains, most commonly energy, physical appearance, general health, and physical strength, as well as hydration and gut health (identified from open-text responses during coding; full taxonomy in **eMethods 1**). Harm-related information was coded when presentations mentioned side effects, overdose risks, drug interactions, lack of testing, contamination risks, or unnecessary use.

### Statistical Analysis

Descriptive statistics were used to summarize influencer characteristics, supplement types, advertising and disclosure, persuasive strategies, and benefit and harm communication. The unit of analysis was the individual supplement presentation (n = 1,120). Because multiple supplement presentations could originate from the same influencer account, mixed-effects logistic regression models with random intercepts for influencer accounts were used to examine associations with personal experience narratives and benefit claims. Models examining influencer characteristics included health-related credentials, follower reach category, and gender as predictors. To examine associations with supplement type, separate models were fitted for sports nutrition, micronutrients, stimulants, miscellaneous supplements, and presentations for which no specific ingredient was identified because these categories were not mutually exclusive. These models were adjusted for influencer health-related credentials, follower reach category, and gender. Odds ratios (ORs) with 95% CIs are reported. There were no missing data for variables included in the regression analyses; therefore, no missing-data procedures were required.

Promotional and negative tone, evidence cues, and harm communication were summarized descriptively because their low event frequencies and concentration within a small number of influencer accounts resulted in unstable mixed-effects model estimates. Analyses were exploratory, and P values were not adjusted for multiple comparisons. All tests were 2-sided, with P<.05 considered statistically significant. Analyses were conducted in R version 4.3.0 (R Foundation for Statistical Computing).

## Results

### Sample Characteristics

A total of 1,120 individual dietary supplement presentations were identified across 727 Instagram posts published between 2022 and 2025 by 84 influencers (**Table 1**). Most influencer accounts had large audiences, with 61 accounts (72.6%) classified as mega influencers (≥300,000 followers). The median number of followers was 650,500 (IQR, 2,289,500). Posts were primarily in English (374 [51.4%]) or German (290 [39.9%]) and differed widely in user engagement (likes: 0-15,900,000; median, 7,875; mean, 99,076; comments: 0-121,000; median, 91; mean, 1,009).

**Table 1.** Characteristics of Influencers, Posts, and Supplement Presentations.

| **Characteristic** | **No. (%)** |
|---|---|
| **Influencers, total** | 84 |
| **Posts, total** | 727 |
| **Supplement presentations, total** | 1,120 |
| **Influencer-level characteristics (n = 84)** |  |
| **Health credentials** |  |
| Health professional<sup>a</sup> | 5 (6.0) |
| Non–health professional | 79 (94.0) |
| **Follower reach** |  |
| Micro (<20,000) | 4 (4.8) |
| Meso (20,000–<100,000) | 5 (6.0) |
| Macro (100,000–<300,000) | 14 (16.7) |
| Mega (≥300,000) | 61 (72.6) |
| **Country** |  |
| Germany | 31 (36.9) |
| USA | 16 (19.0) |
| Austria | 8 (9.5) |
| Other | 29 (34.5) |
| **Verification status** |  |
| Verified | 69 (82.1) |
| Not verified | 15 (17.9) |
| **Gender** |  |
| Female | 36 (42.9) |
| Male | 48 (57.1) |
| Non-binary | 0 (0.0) |
| **Post-level characteristics (n = 727)** |  |
| **Language** |  |
| English | 374 (51.4) |
| German | 290 (39.9) |
| Mixed/Other | 63 (8.7) |
| **Product-level characteristics (n = 1,120)** |  |
| **Commercial presentation** |  |
| Advertising present | 1,088 (97.1) |
| Disclosure present | 286 (26.3 within advertised) |
| Disclosure absent | 802 (73.7 within advertised) |
| Advertising absent<sup>b</sup> | 32 (2.9) |
| **Tone of product evaluation** |  |
| Promotional | 1,092 (97.5) |
| Negative | 27 (2.4) |
| Neutral | 1 (0.1) |

<sup>a</sup>Health professionals were defined as influencers who self-reported formal health-related credentials and included two physicians, one psychiatric nurse, one nutrition scientist, and one dietitian. <sup>b</sup>Advertising absent refers to posts discussing dietary supplements without commercial endorsement.

### Types of Supplements Presented

Protein was the most frequently presented ingredient (463 [41.3%]), followed by caffeine (156 [13.9%]) and creatine (71 [6.3%]). In contrast, micronutrients were less frequently represented, including (multi-)vitamins (67 [6.0%]) and (multi-)minerals (61 [5.4%]) (**Table 2**). When grouped into analytic categories, sports nutrition supplements accounted for 566 (50.5%), followed by micronutrients (103 [9.2%]), stimulant products (156 [13.9%]), miscellaneous ingredients (173 [15.4%]), and presentations with no specific ingredient identified (168 [15.0%]). Furthermore, 107 distinct brands were identified across all presentations. A detailed list of identified ingredients is provided in **eTable 2**, product forms in **eTable 3**, and brand distribution in **eTable 4**.

**Table 2.** Identified Ingredients of Supplements and Aggregated Categories (n = 1,120)

| **Supplement category** | **No. (%)** |
|---|---|
| **Sports nutrition ingredients** | **566 (50.5)** |
| Proteins | 463 (41.3) |
| Creatine | 71 (6.3) |
| Amino acids | 32 (2.9) |
| **Micronutrient ingredients** | **103 (9.2)** |
| (Multi-)vitamins | 67 (6.0) |
| (Multi-)minerals | 61 (5.4) |
| **Stimulant ingredients** | **156 (13.9)** |
| Caffeine | 156 (13.9) |
| **Miscellaneous** | **173 (15.4)** |
| Herbals | 29 (2.6) |
| Collagen | 28 (2.5) |
| Pre- and probiotics | 20 (1.8) |
| Fatty acids (omega-3/6) | 20 (1.8) |
| Antioxidants | 4 (0.4) |
| Testosterone boosters | 4 (0.4) |
| Melatonin/sleep aids | 1 (0.1) |
| Other | 85 (7.6) |
| **No specific ingredient identified** | 168 (15.0) |

*Note: Ingredient categories are not mutually exclusive; percentages do not sum to 100%.*

### Overall Tone, Strategies, and Advertising Disclosure

Among 1,120 supplement presentations, nearly all involved advertising (1,088 [97.1%]) and had an overall positive tone (1,092 [97.5%]). Among the advertisements, disclosure was absent in 802 cases (73.7%). A negative or warning tone was observed in 27 (2.4%) and neutral or balanced communication in 1 (0.1%). Persuasive strategies were predominantly experiential, with personal experience narratives used in 458 supplement presentations (40.9%). Furthermore, actual consumption occurred in 244 cases (21.8%), while holding or using the product occurred in 345 presentations (30.8%) (**Table 3**).

**Table 3.** Content Characteristics of Dietary Supplement Presentations (n = 1,120)

| **Content characteristic** | **No. (%)** |
|---|---|
| **Benefit and risk information** |  |
| At least one benefit mentioned | 269 (24.0) |
| At least one potential harm mentioned | 26 (2.3) |
| **Behavioral modeling** |  |
| Holding or using the product | 345 (30.8) |
| Consuming the product | 244 (21.8) |
| Using for recipe (not consuming) | 123 (11.0) |
| **Persuasive strategies** |  |
| Personal experience | 458 (40.9) |
| Evidence cues | 42 (3.8) |

*Note:* Content characteristics are not mutually exclusive.

### Risk and Benefit Communication

Explicit benefits were mentioned in 269 supplement presentations (24.0%), most commonly related to energy, appearance, general health, and strength (**eTable 1**). In contrast, potential risks were mentioned in only 26 (2.3%), most often framing supplements as unnecessary or cautioning against side effects. No presentation mentioned drug interactions or contamination risks.

### Differences Across Supplement and Influencer Types

Communication characteristics varied descriptively between influencers with and without health-related credentials (**Figure 1** and **eTable 5**). Evidence cues were present in 12 of 22 supplement presentations (54.5%) by health professionals compared with 30 of 1098 presentations (2.7%) by non-health professionals, whereas personal experience narratives were less common among health professionals (1 of 22 [4.5%] vs 457 of 1098 [41.6%]). Given the small number of health professionals (n = 5 influencers) and sparse outcome distributions, these differences were not evaluated inferentially for evidence cues, tone, or harms. Adjusted models for personal experience narratives and benefit claims by influencer characteristics are presented in **eTable 7**.

**Figure 1.**
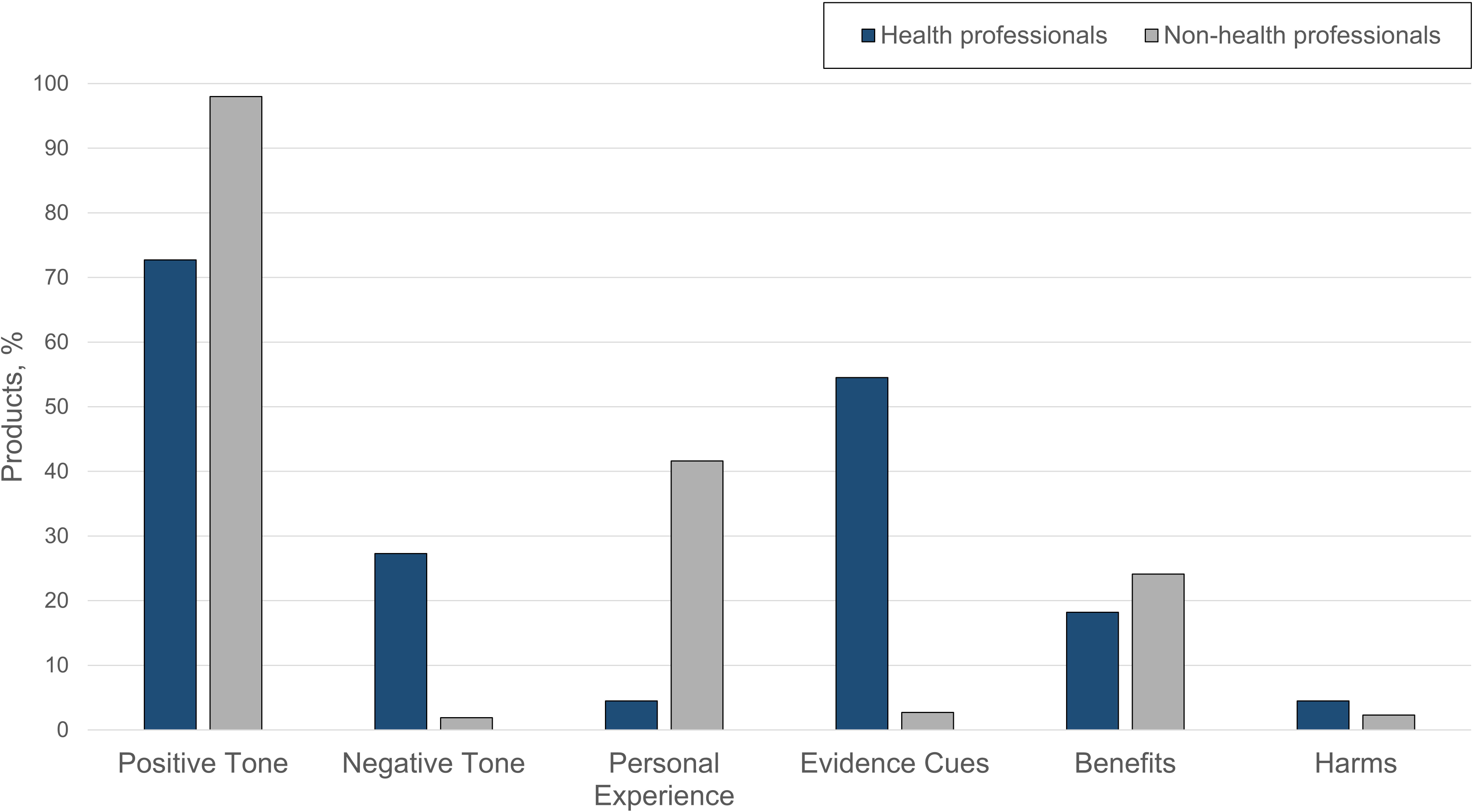
Content Characteristics of Dietary Supplement Presentations by Influencer Type. Percentages are based on supplement presentations by influencers with and without health-related professional credentials.

Communication characteristics also varied descriptively across supplement categories (**Figure 2** and **eTable 6**). In adjusted mixed-effects models (**eTable 8**), personal experience narratives were less common in presentations of stimulants (OR, 0.50; 95% CI, 0.31-0.83; P=.007), with no evidence of associations for the other supplement categories. Benefit claims were less common in presentations of sports nutrition products (OR, 0.33; 95% CI, 0.24-0.47; P<.001) and more common in presentations of micronutrients (OR, 1.88; 95% CI, 1.11-3.19; P=.019), stimulants (OR, 2.38; 95% CI, 1.45-3.90; P<.001), miscellaneous supplements (OR, 1.98; 95% CI, 1.29-3.03; P=.002), and supplements for which no specific ingredient was identified (OR, 1.81; 95% CI, 1.18-2.79; P=.007).

**Figure 2.**
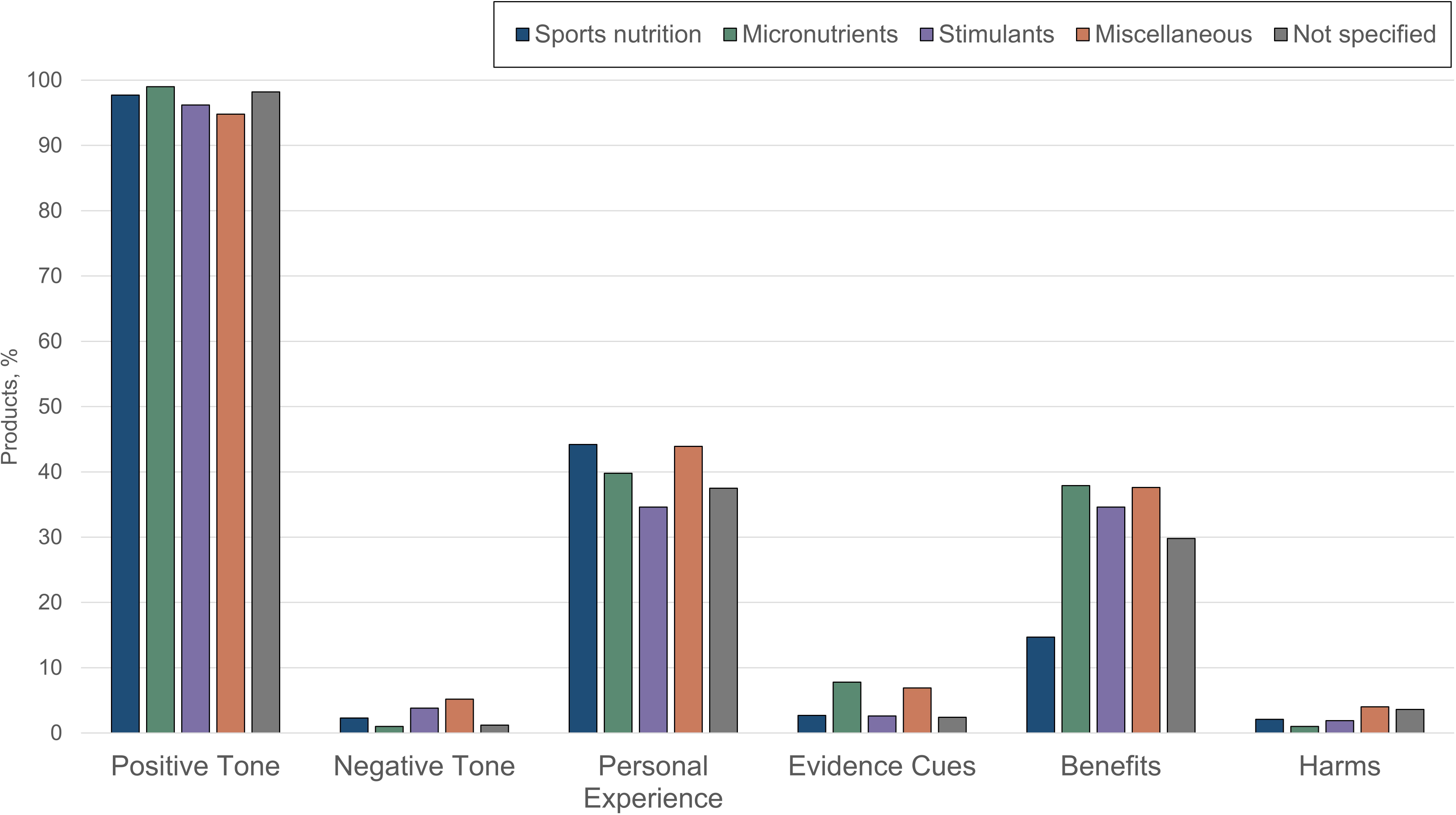
Content Characteristics of Dietary Supplement Presentations by Main Supplement Category. Percentages are based on supplement presentations within each supplement category. Categories are not mutually exclusive.

## Discussion

In this cross-sectional content analysis of Instagram influencers identified through audience-centered sampling with adolescents, dietary supplement presentation was highly commercialized and frequently undisclosed, with a marked imbalance between benefit and risk communication. Only 5 of 84 influencers had health-related professional credentials. Personal experience narratives were common, whereas evidence cues and clinically relevant risk information were rare. Sports nutrition ingredients accounted for approximately half of supplement presentations, but benefit claims were less common in this category than in presentations without sports nutrition ingredients. By contrast, benefit claims were more common in presentations of micronutrients, stimulants, miscellaneous supplements, and supplements for which no specific ingredient was identified. These findings suggest that although benefit framing varies across supplement categories, limited communication of evidence and potential harms is a broader feature of influencer supplement content.

Risk information was rare across supplement categories, with harms mentioned in only 1.0% to 4.0% of presentations within individual categories. This scarcity may be particularly consequential for adolescents, among whom dietary supplement use is common and may occur without clinical guidance.^22^ Sports nutrition supplements, including protein and creatine, were especially prominent in our sample and are widely consumed by adolescents,^23^ raising particular concerns for individuals with pre-existing renal impairment.^24^ Stimulant supplements containing caffeine carry documented cardiovascular risks, with particular concerns for individuals with underlying cardiac conditions.^25^ Herbal ingredients are also associated with clinically relevant safety concerns, including potential hepatotoxicity and interactions with prescription medicines.^26,27^ Yet harm information was uncommon overall, and no presentation mentioned drug interactions or contamination risks.

These findings extend prior research on influencer presentation of health-related products, including prescription drugs^13,14^ and medical tests,^15,16^ which has documented limited disclosure and selective presentation of benefits. For comparison, a recent study found that 33.4% of influencer prescription drug promotions included risk information, while 14.7% of promotions of medical self-tests included such information.^16^ The 2.3% observed for supplement presentations in our study suggests that the imbalance between benefit and risk communication may be particularly pronounced for dietary supplements. One possible contributor is the regulatory distinction between supplements and prescription medicines, which in some jurisdictions are subject to more stringent requirements for communicating risks. However, regulatory requirements vary across countries and platforms, and the present study was not designed to test the effects of regulatory regimes.

The combination of pervasive advertising, limited disclosure, and personal experience narratives may be particularly relevant for adolescent audiences. Commercial messages embedded within fitness, health, and lifestyle content may make persuasive intent difficult to recognize and require adolescents to simultaneously evaluate both the commercial nature and health-related quality of the information. The scarcity of evidence cues and risk information may further limit opportunities to critically assess supplement claims.

These findings have implications for clinical practice, platforms, and regulation. For pediatricians and other clinicians caring for adolescents, asking about supplement use and the social media sources shaping health, fitness, and nutrition decisions may provide opportunities to identify use initiated without professional guidance and discuss potential benefits, harms, and interactions. For platforms, the high prevalence of undisclosed commercial content points to challenges in the implementation and enforcement of existing disclosure requirements. For regulators, the marked imbalance between benefit and risk communication raises the question of whether existing rules governing supplement promotion provide adequate safeguards, particularly for younger audiences in social media environments where commercial messages are embedded within personal health and lifestyle content.

### Strengths and Limitations

Strengths include the audience-centered sampling strategy, which identified influencers through 2 complementary youth-focused approaches: adolescents’ reported encounters with social media accounts and participatory identification and sampling of influencers and their content. This approach provided a sample grounded in social media environments relevant to adolescents rather than researcher-defined lists of health or supplement influencers. The systematically derived dataset of 1,120 supplement presentations enabled detailed product-level analysis. Further strengths include the theory-informed coding scheme, accounting for clustering at the influencer level, and the preregistered and transparent study procedures. One preregistered research question comparing advertised and non-advertised supplement presentations could not be addressed because nearly all presentations were classified as advertising.

Several limitations should be considered. The analysis was restricted to Instagram and may not capture promotional practices on other platforms such as TikTok or YouTube. The sample was limited to German- and English-language content from influencers identified through studies of adolescents in 2 Central European countries, which may limit generalizability to other linguistic, cultural, and geographic contexts. The small number of influencers with health-related credentials limited comparisons between influencers with and without such credentials. Although the audience-centered sampling strategy identified influencers relevant to adolescent social media environments, the study did not establish that individual adolescents were exposed to the specific supplement presentations analyzed. The study captured communicated content but could not assess audience interpretation or behavioral effects. Finally, classification of advertising and disclosure was based on observable content and predefined coding criteria. Underlying commercial agreements between influencers and brands could not be independently verified.

## Conclusions

Dietary supplement presentations by social media influencers identified through audience-centered sampling with adolescents were characterized by pervasive commercialization, limited disclosure, and a marked imbalance between benefit and risk communication. No presentations communicated drug interactions or contamination risks. Benefits were mentioned approximately 10 times as often as harms, and nearly three-quarters of advertisements lacked disclosure. These findings suggest that social media environments relevant to adolescents may provide limited information for recognizing commercial intent and evaluating the potential risks of dietary supplements. Pediatricians and other clinicians caring for adolescents should consider discussing supplement use and social media information sources with their patients. The findings also point to challenges in the implementation and enforcement of existing disclosure requirements and raise the question of whether additional standards are needed to promote more transparent and balanced communication of supplement benefits and risks on social media.

## Data Sharing Statement

The data and analytic code underlying the findings of this study are available at OSF: https://osf.io/rw9pz.

## Author Contributions

Ms Engel and Dr Heiss contributed to the concept and design. Ms Engel, Dr Heiss, Ms Möri, and Ms Vos contributed to acquisition of data. Ms Engel conducted the statistical analysis. All authors contributed to interpretation of the findings and critical revision of the manuscript for important intellectual content. Dr Heiss supervised the study.

## Additional Contributions

We thank Katarina Francisci and Julian Schmidhofer for their assistance with coding. They provided written permission to be named in the manuscript and received no compensation for their contributions.

## Data Access and Responsibility

Ms Engel and Dr Heiss had full access to all the data in the study and take responsibility for the integrity of the data and the accuracy of the data analysis.

## Use of Artificial Intelligence

ChatGPT (GPT-5.6, OpenAI) was used during manuscript preparation in August and September 2026 to assist with formatting, language editing, and developing R code used for the statistical analyses. All AI-assisted outputs were reviewed and verified by the authors, who take full responsibility for the accuracy and integrity of the submitted content.

