## Supplementary material for "Dietary Supplement Promotion by Social Media Influencers: Risk Omission and Limited Disclosure": Online Supplement

This document contains supplemental material cited in the manuscript.

**Contents**

eMethods 1. Codebook

eMethods 2. Audience-Centered Sampling and Source Studies

eMethods 3. Interrater Reliability

eMethods 3a. Interrater Reliability — Round 1 (3 Coders)

eMethods 3b. Interrater Reliability — Round 2 (2 Coders)

eTable 1. Benefits and Harms Explicitly Mentioned in Dietary Supplement Presentations (n = 1,120)

eTable 2. Identified Ingredients of Dietary Supplement Products (n = 1,120 products; multiple ingredients possible per product)

eTable 3. Forms of Dietary Supplement Products (n = 1,120 products)

eTable 4. Brands of Promoted Dietary Supplement Products (n = 1,120 products)

eTable 5. Content Characteristics by Influencer Health Credentials

eTable 6. Content Characteristics by Supplement Category

eTable 7. Adjusted Mixed-Effects Logistic Regression Models for Personal Experience Narratives and Benefit Claims by Influencer Characteristics

eTable 8. Adjusted Mixed-Effects Logistic Regression Models for Personal Experience Narratives and Benefit Claims by Supplement Category

**eMethods 1. Codebook**

The coding scheme was developed using a combined deductive-inductive approach informed by prior literature and refined through three iterative pretesting rounds. The complete codebook is available at the OSF preregistration (https://osf.io/rw9pz). The table below describes all coded variables, their operationalizations, response options, and methodological references.

| **Variable** | **Description** | **Values / Coding** | **Notes / References** |
| --- | --- | --- | --- |
| **A. General data** | | | |
| DATE_CODE | Date the post was coded. | DD.MM.YYYY | — |
| ID | Sequential influencer and post ID. | Numeric (eg, 113, 93) | Pre-allocated in data file. |
| PROFIL_LINK | Full URL of influencer profile. | Text (https://www.instagram.com/…) | Must use standard URL format. |
| **B. Influencer profile** | | | |
| VERIFIED | Profile verified by Instagram? | 0 = No; 1 = Yes | Blue tick next to profile name. |
| GENDER | Gender displayed or perceived. | 1 = Female (she/her); 2 = Male (he/him); 3 = Non-binary (they/them); 0 = Unclear | Code perceived gender unless pronouns explicitly stated. Non-binary = any pronoun except she/her or he/him. |
| AGE | Age if available in profile or via one web search. | Numeric; blank if unclear | famousbirthdays.com may be used. |
| FOLLOWER | Follower count from profile. | Numeric (write out fully: 342000) | Found at top of Instagram profile. |
| COUNTRY | Country of residence. | 1 = Germany; 2 = Austria; 3 = Switzerland; 4 = Other Europe; 5 = USA; 6 = Other | From profile or quick web search. |
| POSTS | Total posts published on account. | Numeric | Found next to follower count. |
| PROFESSION | Any self-reported profession or expertise. | Open text; blank if unavailable | From profile name or description. |
| **C. Post-level variables** | | | |
| POST_LINK | Full URL of the post. | Text | Pre-allocated in data file. |
| DATE | Publication date of post. | DD.MM.YYYY | — |
| CAROUSEL | Number of slides in a carousel post. | 1, 2, 3, … or 0 if not applicable | Enter 0 if single image or video. |
| LANGUAGE | Language(s) of the post. | 0 = Other; 1 = German; 2 = English; 3 = English & German; 4 = German & Other; 5 = English & Other | Includes spoken, caption, and on-screen text. |
| DESCRIPTION | Full text of the post caption. | Open text field | Include all text and hashtags. |
| FORMAT_VIDEO / FORMAT_IMAGE | Format of post content. | 0 = No; 1 = Yes (coded separately for each) | Carousel posts may contain both. |
| LIKES / COMMENTS | Engagement metrics. | Numeric (write out fully) | Often displayed as '320k'. |
| **D. Product-level variables** | | | |
| PRODUCT_NUMBER | Unique product identifier per post. | Influencer ID + sequential number (eg, 66_1, 66_2) | Same product in different flavors counts as one. |
| CLOTHING | Clothing worn while product is shown. | 0 = Product only; 1 = Underwear/Lingerie; 2 = Swimsuit; 3 = Sport/gym; 4 = Casual; 5 = Medical (white coat, scrubs) | Code 0 if no influencer visible. [1,2] |
| TONE | Overall tone regarding the supplement. | 1 = Promotional/positive; 2 = Negative/warning; 3 = Neutral/balanced | If advertisement, code as 1 unless explicitly otherwise. Code 2 if harm discussion outweighs benefits. [3,4] |
| **E. Supplement form** | | | |
| FORM variables (9 binary) | Physical form of the supplement. | 0 = No; 1 = Yes for each: liquid, pill/capsule/tablet, powder, gummies, bar/ball, drops, spray, gel, other (open text) | Energy drinks = liquid; protein shakes from powder = powder only. Multiple forms possible per product. [3] |
| **F. Supplement ingredients** | | | |
| SUPPL_INGREDIENTS (14 binary + other) | Ingredients identifiable from the post. | Binary (0/1) for each of 14 categories: any vitamin present, any mineral present, caffeine, protein, creatine, amino acids (BCAA/EAA), herbals, omega-3/6, melatonin/sleep aids, collagen, antioxidants, testosterone, probiotics, prebiotics; plus open text. Individual vitamin and mineral subtypes were not analyzed separately. | Code only ingredients explicitly shown or named. Energy drinks always coded for caffeine unless labeled caffeine-free. [5,6] |
| **G. Advertising and disclosure** | | | |
| ADVERTISEMENT | Is the product advertised (ie, is there a financial interest behind the product mention)? | 0 = No; 1 = Yes | A product presented without financial interest, for example negatively in a reaction video, was not classified as advertising. |
| DISCLOSURE | Is the commercial relationship disclosed? (only if ADVERTISEMENT = 1) | 0 = No; 1 = Yes | Disclosure = verbal, textual, or visual indicator (eg, #ad, #sponsored, 'Werbung', platform-native badge). Brand/product tagging alone is NOT sufficient disclosure. [7] |
| DISCOUNT | Discount or promotional code offered? | 0 = No; 1 = Yes | Includes variant spellings (eg, c0de). [8] |
| **H. Persuasive strategies** | | | |
| EVIDENCE | Any reference to empirical evidence? | 0 = No; 1 = Yes | Eg, screenshots of studies, 'studies have shown…'. [9] |
| EXPERTISE | Any reference to professional credentials? | 0 = No; 1 = Yes | Eg, white coat, stethoscope, 'as a doctor I recommend…'. [10,11] |
| EXPERIENCE | Positive personal experience reported? | 0 = No; 1 = Yes | Includes consuming product in post. [11,12] |
| HYPERBOLIC | Hyperbolic language or exaggerated visuals? | 0 = No; 1 = Yes | Eg, 'miracle product', 'instant results'. [13] |
| HUMOR | Humor present? | 0 = No; 1 = Yes | Unusual/surprising situation, sarcasm, jokes, or self-deprecation. [12,14] |
| TARGETGROUP / TARGETGROUP_SPECIFIC | Specific target group addressed? | 0 = No; 1 = Yes; open text for specific group | Not coded if benefits merely imply a target group. |
| INTERACTION | How does influencer interact with product? | 0 = No interaction; 1 = Holding/presenting but not consuming; 2 = Using for recipe but not consuming end product; 3 = Consuming the product or recipe containing it | [3] |
| **I. Calls to action** | | | |
| CALL_BUY / CALL_INFO / CALL_CONSULT / CALL_SELFTEST | Calls to action present? | 0 = No; 1 = Yes for each | CALL_CONSULT = encourage to consult health professional. CALL_SELFTEST = encourage at-home self-testing. [10,15] |
| **J. Health content: benefits and harms** | | | |
| HEALTH CLAIMS & BENEFITS | Health benefit claims present? | 0 = No; 1 = Yes. Subcategories: 1 = Cure disease; 2 = Prevention; 3 = Appearance; 4 = Mental wellbeing; 5 = Cognitive enhancement; 6 = Pain relief; 7 = Strength; 8 = Energy; 9 = Immune boost; 10 = Recovery; 11 = General health; Other = open text | [16] |
| HARMS / RISKS | Harms, risks, or negative aspects mentioned? | 0 = No; 1 = Yes. Subcategories: 1 = Side effects; 2 = Overdose; 3 = Drug interaction; 4 = Untested; 5 = Contamination; 6 = Unnecessary; 7 = No replacement for whole foods; Other = open text | [10] |
| DOSAGE | Usage or dosing instructions provided? | 0 = No; 1 = Yes | Eg, '2 tablespoons per day', '1 pill weekly'. |

Abbreviations: BCAA = branched-chain amino acids; EAA = essential amino acids. Energy drinks were coded as caffeine (ingredient code 17) by default as caffeine is a regulated inherent ingredient, unless the product is explicitly labeled caffeine-free. Supplement analytic categories (sport nutrition, stimulant, micronutrient, miscellaneous, unspecified) were derived by grouping individual ingredient codes post-hoc as described in the Methods section.

**eMethods 2. Audience-Centered Sampling and Source Studies**

Influencer accounts were derived from 2 independent youth-focused studies in Switzerland and Austria using complementary approaches to identify social media accounts relevant to adolescents.

In the Swiss study, 1392 adolescents from 12 secondary and vocational schools participated in the third survey wave. Participants were first asked how often they encountered social media posts intended to motivate a healthier lifestyle, such as through physical activity or healthy eating. Those reporting such content were subsequently asked to nominate specific accounts from which they frequently encountered these messages. For Instagram, 1125 adolescents received the account-nomination question and 339 provided responses, yielding 210 accounts belonging to individual creators. Further details of the study design and data collection are reported in Möri et al.[1]

In the Austrian study, a participatory citizen science approach was used in which adolescents acted as citizen youth scientists and contributed to data collection and categorization. The study included 336 adolescents aged 12 to 19 years from 19 classes across 16 Austrian schools, including schools in both rural and urban areas. Teachers and students received standardized information materials and a video tutorial on the research process and data collection. The data collection tool was pretested with a middle school class of 18 students and 2 teachers. During a designated data collection week between April and May 2023, participating classes identified influencers they considered most important after consulting classmates in a plenary session. They then documented at least 5 publicly available health-related social media posts per selected influencer using a standardized online data collection tool. Data collection covered Instagram, TikTok, and YouTube. Overall, adolescents documented 2147 posts from 255 influencer profiles, 95.3% of which had an Instagram presence. Full details of the study design, sampling, and data collection are reported in Gell et al.[2]

The account pools from both studies were subsequently combined for the present study. Between August and September 2024, duplicates were removed, non-Instagram accounts were excluded, and accounts were screened for health-related content, including dietary supplements. This audience-centered approach therefore identified influencers based on their relevance to adolescent audiences rather than researcher-defined lists of health or supplement influencers.

***References for eMethods 2***

1. Möri M, Tribelhorn L, Friemel TN. The influence of presumed social media influence: a longitudinal analysis of social media influence on adolescents’ physical activity. *SCM Stud Commun Media*. 2025;26. doi:10.31234/osf.io/7zteu_v1

2. Gell S, Engel E, Karsay K, Heiss R. Uncovering social media influencers’ health posts: a youth-driven mixed method study. *medRxiv*. Preprint posted online October 14, 2026. doi:10.64898/2026.10.13.26365375.

**eMethods 3. Interrater Reliability**

Interrater reliability was assessed iteratively during coder training and refinement of the coding scheme. An initial reliability assessment was conducted by three coders on 42 supplement presentations. For variables that did not demonstrate adequate reliability, coding definitions were refined and/or categories were collapsed where appropriate. A second independent reliability assessment was then conducted by two coders on 113 supplement presentations using the revised coding scheme.

**eMethods 3a. Interrater Reliability — Round 1 (3 Coders, n = 42 products; variables with α ≥ 0.80 or 100% percentage agreement)**

| **Variable** | **α** | **PA, %** |
| --- | --- | --- |
| **Persuasive Strategies** | | |
| Evidence cues | 0.897 | 98.3 |
| **Supplement Form** | | |
| Powder | 0.873 | 93.7 |
| Gummies | 1.000 | 100.0 |
| Bar/Ball | 1.000 | 100.0 |
| Drops | NA^a^ | 100.0 |
| Spray | NA^a^ | 100.0^b^ |
| Gel | 1.000 | 100.0^b^ |
| **Supplement Ingredients** | | |
| Protein | 0.963 | 98.4 |
| Creatine | 0.920 | 98.4 |
| Amino acids | NA^a^ | 100.0 |
| Collagen | 1.000 | 100.0 |
| Omega-3/6 fatty acids | 0.900 | 98.4 |
| Melatonin | NA^a^ | 100.0 |
| Testosterone boosters | NA^a^ | 100.0^b^ |

Abbreviations: α, Krippendorff alpha; PA, percentage agreement.

a Statistic not calculable because coders used only one category for all items; PA = 100% indicates perfect agreement.

b Based on n = 17 items (Rounds 2 and 3 only).

**eMethods 3b. Interrater Reliability — Round 2 (2 Coders, n = 113 products)**

| **Variable** | **κ** | **PA, %** |
| --- | --- | --- |
| **Post-Level Variables** | | |
| Tone^a^ | 1.000 | 100.0 |
| Advertisement present | 1.000 | 100.0 |
| Disclosure present | 0.868 | 93.8 |
| Visual interaction with product^a^ | 0.955 | 94.7 |
| **Persuasive Strategies** | | |
| Personal experience narrative | 0.805 | 90.3 |
| **Benefit and Harm Claims** | | |
| Benefit claimed | 0.831 | 92.0 |
| Harm disclosed | 1.000 | 100.0 |
| **Supplement Form** | | |
| Liquid | 1.000 | 100.0 |
| Pill | 1.000 | 100.0 |
| **Supplement Ingredients** | | |
| Any vitamin present | 0.958 | 99.1 |
| Any mineral present | 1.000 | 100.0 |
| Caffeine | 1.000 | 100.0 |
| Herbal ingredients | 1.000 | 100.0 |
| Antioxidants | 1.000 | 100.0 |

Abbreviations: κ, Cohen kappa; PA, percentage agreement.

a Ordinal variable; weighted kappa (linear weights) reported.

**eTable 1. Benefits and Harms Explicitly Mentioned in Dietary Supplement Presentations (*n* = 1,120)**

| **Category** | **No. (%)** |
| --- | --- |
| **Benefits (*n* = 269 presentations with ≥1 benefit mentioned; 24.0% of total)** | |
| Energy | 76 (6.8%) |
| Physical appearance | 64 (5.7%) |
| General health | 47 (4.2%) |
| Physical strength | 40 (3.6%) |
| Recovery | 12 (1.1%) |
| Cognitive enhancement | 12 (1.1%) |
| Hydration | 27 (2.4%) |
| Gut health | 14 (1.2%) |
| Immune boost | 6 (0.5%) |
| Mental wellbeing | 7 (0.6%) |
| Disease prevention | 6 (0.5%) |
| Pain relief/management | 1 (0.1%) |
| Disease cure | 6 (0.5%) |
| Othera | 56 (5.0%) |
| **Harms (n = 26 presentations with ≥1 harm mentioned; 2.3% of total)** | |
| Unnecessary | 22 (2.0%) |
| Side effects | 6 (0.5%) |
| No replacement for healthy diet | 6 (0.5%) |
| Overdose risk | 3 (0.3%) |
| Drug interaction with other medication | 0 (0.0%) |
| Untested/unproven | 0 (0.0%) |
| Contamination risk | 0 (0.0%) |

Percentages are of total supplement presentations (*n* = 1,120). Because a single presentation could mention multiple benefit or harm categories, counts within each section do not sum to section totals. ^a^ “Other” benefits include heterogeneous claims not mapping onto predefined categories, most commonly weight loss/fat loss (n = 5), sleep (n = 2), sugar-free/clean formulation (n = 2), blood flow, hitting nutrition goals, supporting training, reducing inflammation, blood sugar control, increased testosterone, and facilitating fatty acid transport. Some entries in this field were promotional or taste-related statements (eg, “tastes great,” brand-loyalty phrases) rather than substantive health, performance, or appearance claims; these were retained in the free-text field but did not independently drive the benefit-mentioned classification beyond what other coded variables already captured.

**eTable 2. Identified Ingredients of Dietary Supplement Products (*n* = 1,120 products; multiple ingredients possible per product)**

| **Ingredient** | **No. (% of products)** |
| --- | --- |
| **Sport nutrition ingredients** | |
| Protein | 463 (41.3%) |
| Creatine | 71 (6.3%) |
| Amino acids (BCAA, EAA, and others) | 32 (2.9%) |
| **Stimulant ingredients** | |
| Caffeine | 156 (13.9%) |
| **Micronutrient ingredients** | |
| (Multi-)vitamins | 67 (6.0%) |
| (Multi-)minerals | 61 (5.4%) |
| **Miscellaneous ingredients** | |
| Herbal ingredients (ashwagandha, maca, ginseng, and others) | 29 (2.6%) |
| Collagen/collagen peptides | 28 (2.5%) |
| Omega-3/6 fatty acids | 20 (1.8%) |
| Prebiotics | 11 (1.0%) |
| Probiotics | 10 (0.9%) |
| Melatonin/sleep aids | 1 (0.1%) |
| Antioxidants/longevity compounds | 4 (0.4%) |
| Testosterone boosters | 4 (0.4%) |
| **No specific ingredient identifiable** | |
| No ingredient identifiable | 168 (15.0%) |

Percentages are of total supplement presentations (*n* = 1,120). Because individual products can contain multiple ingredients, percentages do not sum to 100%. Ingredients are grouped by the analytic categories used in the main analysis. BCAA = branched-chain amino acids; EAA = essential amino acids.

**eTable 3. Forms of Dietary Supplement Products (*n* = 1,120 products)**

| **Form** | **No. (%)** |
| --- | --- |
| Powder | 660 (58.9%) |
| Liquid (ready-to-drink) | 194 (17.3%) |
| Pill/capsule/tablet | 131 (11.7%) |
| Bar/ball | 90 (8.0%) |
| Other | 16 (1.4%) |
| No form identifiable | 4 (0.4%) |
| Drops | 7 (0.6%) |
| Spray | 7 (0.6%) |
| Gel | 6 (0.5%) |
| Gummies | 5 (0.4%) |

**eTable 4. Brands of Promoted Dietary Supplement Products (*n* = 1,120 products)**

| **Brand** | **No. (%)** |
| --- | --- |
| ESN | 191 (17.1%) |
| More Nutrition | 156 (13.9%) |
| Bloom Supps | 77 (6.9%) |
| TC Nutrition | 58 (5.2%) |
| Ryse | 54 (4.8%) |
| Myprotein | 44 (3.9%) |
| WomensBest | 39 (3.5%) |
| Foodspring | 33 (2.9%) |
| MYVEGAN | 31 (2.8%) |
| EVO | 24 (2.1%) |
| Red Bull | 21 (1.9%) |
| Prozis | 21 (1.9%) |
| EVO Sports Fuel | 19 (1.7%) |
| Celsius | 18 (1.6%) |
| Transparent Labs | 18 (1.6%) |
| Lean.foods | 18 (1.6%) |
| Onwards Nutrition | 18 (1.6%) |
| Other (90 additional brands, each n < 18) | 265 (23.7%) |
| No brand identifiable | 15 (1.3%) |

A total of 107 distinct brands were identified after normalizing spelling/capitalization variants; the 17 most frequent are shown individually and the remaining 90 (each n < 18) are grouped as “Other.” Percentages are of total supplement presentations (*n* = 1,120).

**eTable 5. Content Characteristics by Influencer Health Credentials**

| **Content characteristic** | **Health professionals, No./total (%)** | **Non-health professionals, No./total (%)** |
| --- | --- | --- |
| Positive tone | 16/22 (72.7) | 1076/1098 (98.0) |
| Negative tone | 6/22 (27.3) | 21/1098 (1.9) |
| Personal experience | 1/22 (4.5) | 457/1098 (41.6) |
| Evidence cues | 12/22 (54.5) | 30/1098 (2.7) |
| Benefits mentioned | 4/22 (18.2) | 265/1098 (24.1) |
| Harms mentioned | 1/22 (4.5) | 25/1098 (2.3) |

Values are No./total (%). Health professionals were influencers with self-reported formal health-related credentials (n=5 influencers; 22 supplement presentations). Comparisons are descriptive. Positive and negative tone, evidence cues, and harms were not modeled inferentially because low event frequencies and concentration within a small number of influencer accounts resulted in unstable mixed-effects model estimates.

**eTable 6. Content Characteristics by Supplement Category**

| **Content characteristic** | **Sports nutrition (n=566), %** | **Micronutrients (n=103), %** | **Stimulants (n=156), %** | **Miscellaneous (n=173), %** | **Not specified (n=168), %** |
| --- | --- | --- | --- | --- | --- |
| Positive tone | 97.7 | 99.0 | 96.2 | 94.8 | 98.2 |
| Negative tone | 2.3 | 1.0 | 3.8 | 5.2 | 1.2 |
| Personal experience | 44.2 | 39.8 | 34.6 | 43.9 | 37.5 |
| Evidence cues | 2.7 | 7.8 | 2.6 | 6.9 | 2.4 |
| Benefits mentioned | 14.7 | 37.9 | 34.6 | 37.6 | 29.8 |
| Harms mentioned | 2.1 | 1.0 | 1.9 | 4.0 | 3.6 |

Values are percentages of supplement presentations within each category. Supplement categories were not mutually exclusive; therefore, presentations could contribute to more than 1 category. “Not specified” indicates presentations for which no specific supplement ingredient was identified.

**eTable 7. Adjusted Mixed-Effects Logistic Regression Models for Personal Experience Narratives and Benefit Claims by Influencer Characteristics**

| **Predictor** | **Personal experience,**  **OR (95% CI)** | **P value** | **Benefits mentioned,**  **OR (95% CI)** | **P value** |
| --- | --- | --- | --- | --- |
| Health professional | 0.11 (0.01–1.52) | .100 | 0.49 (0.07–3.32) | .463 |
| Follower reach |  |  |  |  |
| Micro | 1 [Reference] | — | 1 [Reference] | — |
| Meso | 1.50 (0.18–12.73) | .709 | 2.05 (0.28–14.87) | .476 |
| Macro | 1.02 (0.18–5.70) | .984 | 1.89 (0.40–9.04) | .425 |
| Mega | 2.49 (0.52–11.81) | .252 | 2.27 (0.55–9.35) | .255 |
| Gender |  |  |  |  |
| Female | 1 [Reference] | — | 1 [Reference] | — |
| Male | 0.32 (0.16–0.65) | .002 | 1.14 (0.59–2.20) | .690 |

OR indicates odds ratio; CI, confidence interval. Mixed-effects logistic regression models included a random intercept for influencer account. Models included health-related credentials, follower category, and gender simultaneously. Reference categories were non-health professional, micro follower reach, and female gender. Analyses were exploratory; P values were 2-sided and were not adjusted for multiple comparisons.

**eTable 8. Adjusted Mixed-Effects Logistic Regression Models for Personal Experience Narratives and Benefit Claims by Supplement Category**

| **Supplement category** | **Personal experience,**  **OR (95% CI)** | **P value** | **Benefits mentioned,**  **OR (95% CI)** | **P value** |
| --- | --- | --- | --- | --- |
| Sports nutrition | 1.06 (0.77–1.45) | .737 | 0.33 (0.24–0.47) | <.001 |
| Micronutrients | 1.34 (0.78–2.28) | .289 | 1.88 (1.11–3.19) | .019 |
| Stimulants | 0.50 (0.31–0.83) | .007 | 2.38 (1.45–3.90) | <.001 |
| Miscellaneous | 1.34 (0.88–2.06) | .176 | 1.98 (1.29–3.03) | .002 |
| Not specified | 1.06 (0.69–1.65) | .786 | 1.81 (1.18–2.79) | .007 |

OR indicates odds ratio; CI, confidence interval. Separate mixed-effects logistic regression models were fitted for each supplement category because categories were not mutually exclusive. The table reports the coefficient for the supplement category from each model. Each model included a random intercept for influencer account and was adjusted for health-related credentials, follower category, and gender. For each supplement category, the reference group was supplement presentations without that category. Analyses were exploratory; P values were 2-sided and were not adjusted for multiple comparisons.
